# Validation of Segmental Bioelectrical Impedance Analysis Compared with Dual Energy X-Ray Absorptiometry to Measure Body Composition in Patients with Obesity-related Heart Failure with Preserved Ejection Fraction

**DOI:** 10.1101/2024.11.20.24317675

**Authors:** Hannah Salmons, Syed Imran Ahmed, Hayley Billingsley, Alexander Reavey-Cantwell, Roshanak Markley, Michele Golino, Marco Giuseppe Del Buono, Juan Ignacio Damonte, Sebastian Pinel, R. Lee Franco, Antonio Abbate, Carrie P. Earthman, Salvatore Carbone

**Affiliations:** Department of Kinesiology & Health Sciences, College of Humanities & Sciences, Virginia Commonwealth University, Richmond, Virginia; Pauley Heart Center, Virginia Commonwealth University, Richmond, Virginia, United States; Robert M. Berne Cardiovascular Research Center, and Division of Cardiology, University of Virginia, Charlottesville, Virginia, United States; Department of Cardiovascular and Pulmonary Sciences, Catholic University of the Sacred Heart, Rome, Italy; Department of Cardiovascular Medicine, Fondazione Policlinico Universitario A. Gemelli IRCCS, Rome, Italy; Hospital Universitario Austral, Pilar, Argentina; Hospital Italiano de Buenos Aires, Ciudad Autónoma de Buenos Aires, Argentina; Department of Health Behavior and Nutrition Sciences, University of Delaware, Newark, DE

**Keywords:** Heart Failure, HFpEF, Obesity, Body Composition, BIA, DXA

## Abstract

**Background:** Appendicular lean mass index (ALMI), a term used to describe appendicular lean soft tissue measured by dual-energy X-ray absorptiometry (DXA), is a major determinant of cardiorespiratory fitness in patients with obesity-related heart failure with preserved ejection fraction (HFpEF). Moreover, ALMI can be used to diagnose sarcopenia and sarcopenic obesity in this population. DXA is a reference standard for assessing body composition, however, segmental single-frequency bioelectrical impedance analysis (SF-BIA) could offer a more accessible, portable, cost-effective, and radiation-free alternative. To validate segmental SF-BIA for body composition analysis in patients with HFpEF and obesity, with a focus on ALMI.

**Methods:** We analyzed 62 consecutive euvolemic patients with persistent obesity-related HFpEF (83.8% female, 60.8± 2.8 years of age). We used both DXA and segmental SF-BIA to measure ALMI and appendicular fat mass index (AFMI), fat mass (FM), fat-free mass (FFM) in kg and as % of body weight, FM index, and FFM index. Correlations were assessed using Pearson’s coefficients and Bland-Altman plots, while linear regression was performed to evaluate proportional bias.

**Results:** Strong, statistically significant correlations were found between BIA and DXA for ALMI (r=0.897), AFMI (r=0.864), FM (r=0.968), FM% (r=0.867), FFM (r=0.954), and FFM% (r=0.852), FM index (r=0.97), and FFM index (r=0.88) (all p<0.001). The Bland-Altman analysis demonstrated agreement between methods and linear regression indicated no significant proportional bias for all parameters, except for AFMI.

**Conclusions:** Segmental SF-BIA-measured body composition shows strong correlations, appropriate agreements, and no proportional bias compared to DXA. Considering the central role of body composition and particularly of ALMI in patients with obesity-related HFpEF, when DXA is not readily available or contraindicated, segmental SF-BIA should be considered in this population.

## INTRODUCTION

Patients with obesity-related heart failure (HF) with preserved ejection fraction (HFpEF) are characterized by excess fat mass (FM) that negatively impacts cardiorespiratory fitness (CRF) and quality of life.^1-5^ The exclusive reliance on body mass index (BMI) may lead to misclassification of these individuals, whereas those with higher BMI, but with greater lean mass (LM, defined as bone-free fat-free mass, FFM), may present a more favorable CRF.^6, 7^ A recent meta-analysis of 27 studies confirmed that reduced CRF measured as peak oxygen consumption (VO_2 peak_) in patients with HF is heavily driven by reduced whole-body LM and appendicular LM.^8^ Moreover, LM, but not FM, is an independent predictor of long-term survival in patients with HFpEF.^9^ When reduced LM is paired with excess adiposity characteristic of obesity (i.e., sarcopenic obesity), prognosis is further worsened in patients with HFpEF.^6, 10-12^ Taken together, this substantiates the limitations of BMI, which does not allow for differentiation between body composition compartments. Importantly, while FFM and LM are often used interchangeably, they provide information on different body composition compartments, with LM being the best surrogate to estimate skeletal muscle mass given that it excludes bone mineral content from FFM (**Figure 1**).

**Figure 1.**
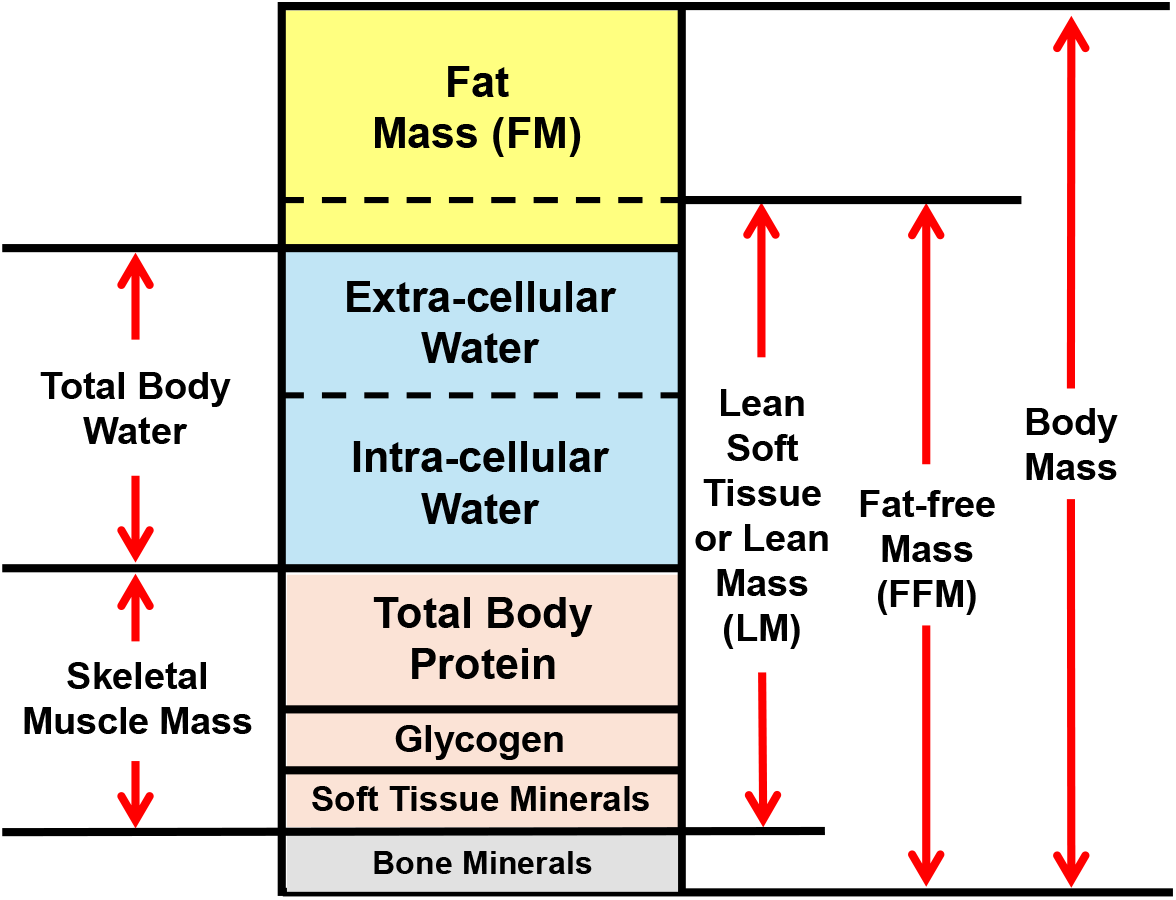
Schematic of body composition compartments that includes the differentiation between fat-free mass, lean mass (or lean soft tissue), and skeletal muscle mass. The sum of lean mass of arms and legs describes appendicular lean mass (ALM). Lean mass divided by height^2^, allows to calculate ALM index (ALMI, kg/m^2^).

Considering the growing use and efficacy of medications for chronic weight management in patients with obesity-related HFpEF,^13^ which results in a reduction of FM and LM, assessing body composition in this population is essential.^14, 15^ The reduction in LM potentially leads to an increased long-term risk for sarcopenia and sarcopenic obesity.^16, 17^

Despite the well-established role of body composition in patients with obesity-related HFpEF, its assessment outside of clinical research is rarely implemented due to the challenges associated with the available tools that can accurately estimate FM and LM compartments. Dual-energy X-ray absorptiometry (DXA) is one of the accepted reference methods for assessing whole-and segmental body composition and diagnosing sarcopenia and sarcopenic obesity.

DXA-measured appendicular lean soft tissue, which is the bone-free FFM of the arms and legs, is the same compartment we refer to as “appendicular lean mass (ALM)”. Although body composition analysis can improve HFpEF risk stratification, there are several barriers to routine implementation of DXA in the clinical setting such as cost, time, weight limit, radiation exposure, lack of portability, and need for skilled personnel for the segmental analysis.^18, 19^ This highlights the need to identify alternative tools to routinely assess whole- and segmental body composition in this population.

Segmental single-frequency bioelectrical impedance analysis (SF-BIA) is a non-invasive tool that provides an estimation of body composition quantity and quality. It may represent a more accessible, portable, and cost-effective method to estimate whole- and segmental body composition, including appendicular LM index (ALMI), without risk of exposure to radiation.

We have previously shown that in patients with obesity-related HFpEF, SF-BIA-derived fat mass index (FM/height^2^) provides a valid alternative to DXA-derived FM index,^4^ and that in patients with HF with reduced ejection fraction, this particular segmental SF-BIA approach is a suitable alternative for assessing ALMI as compared to DXA-assessed ALMI.^6^ This segmental SF-BIA algorithm, however, has not been validated against DXA in patients with obesity-related HFpEF, which would ultimately allow its implementation in both clinical and research practices.^6, 20^

The aim of this study is to validate a segmental SF-BIA method, with a focus on ALMI due to its significant diagnostic and prognostic value in identifying sarcopenia and sarcopenic obesity, against the reference standard DXA in patients with obesity-related HFpEF.

## METHODS

We performed body composition analysis in consecutive adult patients with stable, euvolemic, symptomatic, persistent HFpEF (NYHA class II and III, left ventricular ejection fraction >50% documented in the prior 12 months, no recent hospitalization or changes in medication in the prior month) and obesity, using DXA (Lunar iDXA, encore 2011 software, version 13.60.033, GE HealthCare) and a segmental single-frequency (50 kHz) BIA device (Quantum V, RJL Systems, Clinton Township, Michigan). There were no restrictions on food intake, fluid intake, and physical activity before the visit. For BIA measurements, patients were asked to lie flat on a hospital bed and device-specific electrodes purchased from RJL Systems were placed on both hands and feet in a standard eight-polar configuration. We calculated ALM index (ALMI; kg of LM in limbs/height^2^) and appendicular FM index (AFMI; kg of FM in limbs/height^2^) by both methods. DXA scans provided LM and FM in kg and percentage of body weight (% BW). The segmental SF-BIA device software provide estimates of whole-body and segmental FFM, LM (bone-free fat-free mass, FFM), and FM. The predictive equation used by the segmental SF-BIA for the estimate of right arm, left arm, right leg, and right leg have been described in **Supplemental Table 1**, as provided by the device manufacture. ALM was obtained by summing the LM of both arms and legs, and AFM was obtained by summing the FM of both arms and legs. We further calculated FM index (FMI; kg of FM/height^2^) and FFM index (FFMI; kg of FFM/height^2^). We used transthoracic Doppler echocardiography to measure resting left ventricular ejection fraction and venipuncture to measure glomerular filtration rate, high-sensitivity C-reactive protein, and NT-proBNP.

**Table 1.** Baseline characteristics.

| **Demographic Variable** | **N=62** |
| --- | --- |
| Age in years, $\bar{x} \pm SD$ | 60.8 ($\pm 12.8$) |
| Females, n (%) | 52 (83.8) |
| Black, n (%) | 36 (58) |
| **Functional Class and Comorbidities, n (%)** |  |
| NYHA Class 2 | 52 (83.8) |
| NYHA Class 3 | 10 (16.1) |
| Ischemic Heart Disease | 11 (17.7) |
| Prior Percutaneous Intervention | 8 (12.9) |
| Prior Coronary Artery Bypass Grafting | 3 (4.8) |
| Prior cerebrovascular event | 1 (1.6) |
| Diabetes | 31 (50.0) |
| Hyperlipidemia | 54 (87.1) |
| Tobacco Use | 2 (3.2) |
| Hypertension | 56 (90.3) |
| Peripheral vascular disease | 3 (4.8) |
| Chronic kidney disease | 3 (4.8) |
| Chronic obstructive pulmonary disease | 3 (4.8) |
| Obstructive sleep apnea | 27 (43.5) |
| **Anthropometrics, $\bar{x} \pm SD$** |  |
| Weight (kg) | 101.6±18.8 |
| Height (m) | 1.65±0.079 |
| Body mass index (kg/m<sup>2</sup>) | 36.6±5.4 |
| Waist circumference (cm) | 112.7±13.3 |
| **Vital signs, $\bar{x} \pm SD$** |  |
| Heart rate (bpm) | 71.1±10.8 |
| Systolic blood pressure (mmHg) | 127.7±12.7 |
| Diastolic blood pressure (mmHg) | 68.3±10.4 |
| **Echocardiographic parameters, $\bar{x} \pm SD$** |  |
| Left ventricular ejection fraction (%) | 58.7±4.16 |
| **Biomarkers, $\bar{x} \pm SD$** |  |
| Glomerular filtration rate (mL/min/1.73<sup>2</sup>) | 77.2±19.3 |
| High-sensitivity C-reactive protein (mg/L) | 4.58±5.05 |
| NT-proBNP (pg/mL) | 217.2±307.9 |

$\bar{x}$ , mean; SD, standard deviation; NYHA, New York Heart Association.

Clinical characteristics were presented as number (percentage) or mean±standard deviation (SD). Correlations between DXA and SF-BIA for ALMI (kg/m^2^), AFMI (kg/m^2^), FM (kg), FM%, FFM (kg), FFM%, FMI, and FFMI were assessed using Pearson correlation coefficients. Agreement between the methods was evaluated through Bland-Altman plots and limits of agreement, with coefficients of variation applied to compare BIA measurements against the maximum allowed difference. Simple linear regression was conducted to assess proportional bias in BIA measurements compared to DXA. Statistical analysis was performed using SPSS 28 (IBM Corp., Armonk, New York) and significance was set at p<0.05. All study participants provided written consent.

## RESULTS

### Baseline characteristics

We included sixty-two patients with obesity-related HFpEF, 83.8% self-identified as female and 58% as Black/African American. Mean (±SD) age was 60.8±12.8 years, BMI was 36.6±5.4 kg/m^2^, ejection fraction was 58.7±4.2 %, N-terminal pro-b-type natriuretic peptide was 217.2±307.9 pg/mL, and 52 (83.8%) and 10 (16.2%) patients were classified as NYHA class II and III, respectively (**Table 1**).

### Segmental Body Composition: ALMI and AFMI

ALMI values were strongly correlated between SF-BIA and DXA (r=0.897, p<0.001). Linear regression showed lack of proportional bias (β=0.003, standard error [SE]=0.06, p=0.96), and Bland-Altman analysis indicated limits of agreement from -0.72 to 1.82 kg/m^2^, with all values within the allowed difference range of -3.8 to 3.8 kg/m^2^ (**Figure 2A-B**). Similarly, AFMI values showed a strong correlation (r=0.864, p<0.001), but a significant proportional bias (β=- 0.34, SE=0.068, p<0.001), with limits of agreement from -3.2 to 1.1 kg/m^2^ and the maximum allowed difference to range from -5.0 to 5.0 kg/m^2^. **Table 2** describes the differences of the values between SF-BIA- and DXA-measured segmental body composition compartments.

**Table 2.** Body composition compartments measured with segmental SF-BIA and DXA.

| Body Composition Compartment | SF-BIA | DXA | Difference (BIA-DXA) | 95% CI |  |
| --- | --- | --- | --- | --- | --- |
| | $\bar{x} \pm SD$ | $\bar{x} \pm SD$ | $\bar{x} \pm SD$ | Lower | Upper |
| **ALMI (kg/m<sup>2</sup>)** | 9.32±1.33 | 9.28±1.41 | 0.54±0.64 | 0.3344 | 0.6905 |
| **AFMI (kg/m<sup>2</sup>)** | 6.63±1.53 | 7.68±2.13 | -1.04±1.11 | -1.3898 | -0.7906 |
| **FFM (kg)** | 58.57±10.67 | 55.18±11.23 | 3.38±3.5 | 2.2229 | 4.0742 |
| **FM (kg)** | 44.79±12.19 | 47.34±12.51 | -2.54±3.01 | -3.7097 | -2.0417 |
| **FFM (% BW)** | 57.0±5.81 | 53.59±5.98 | 3.4±3.21 | 2.4194 | 4.1611 |
| **FM (% BW)** | 43.47±6.29 | 45.53±5.86 | -2.54±3.01 | -3.3312 | -1.6752 |
| **FFMI (kg/m<sup>2</sup>)** | 21.28±2.48 | 20.01±2.50 | 1.38±1.34 | 0.8640 | 1.5060 |
| **FMI (kg/m<sup>2</sup>)** | 16.19±4.06 | 17.11±4.14 | -0.91±1.13 | -1.5180 | -0.4220 |

$\bar{x}$ , mean; SD, standard deviation; CI: confidence intervals; BW: body weight; SF-BIA: single frequency segmental bioelectrical impedance analysis; DXA: dual-energy X-ray absorptiometry; ALMI: appendicular lean mass index; AFMI: appendicular fat mass index; FFM: fat-free mass; FM: fat mass; FFMI: fat-free mass index; FMI: fat mass index.

**Figure 2.**
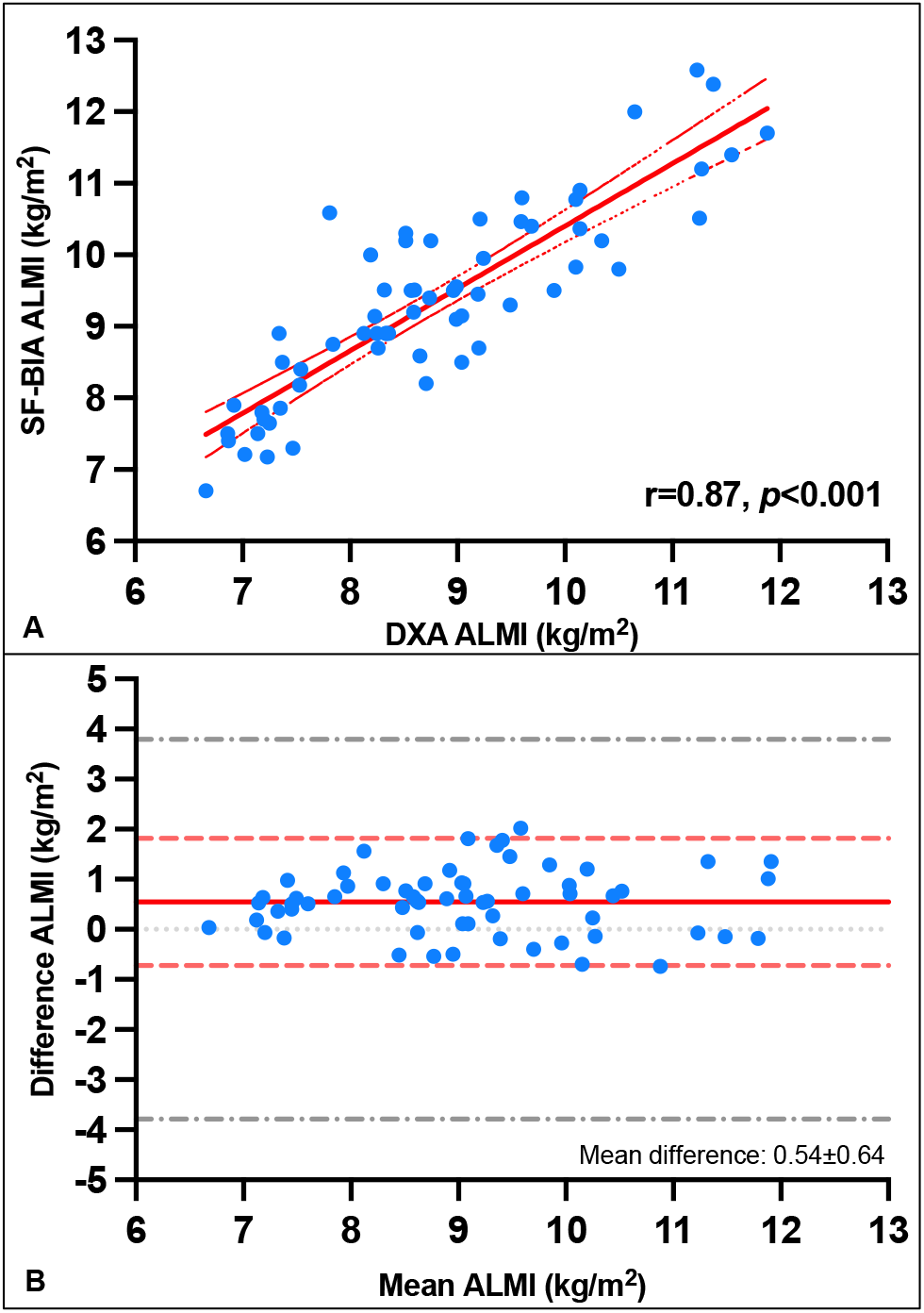
(A) Correlation of appendicular lean mass index (ALMI) (kg/m^2^) between DXA and SF-BIA measurement. (B) Bland-Altman plot with limits of agreements and maximum allowed difference of ALMI (kg/m^2^) between SF-BIA and DXA measurement. Dotted gray lines indicates maximum allowed difference; dotted red lines indicate limits of agreements; solid red line indicates mean difference between SF-BIA and DXA measurements.

### Whole-Body Composition: FM, FFM, FMI, and FFMI

SF-BIA-measured FM (kg) showed a strong, significant correlation with DXA-measured FM (r=0.97, p<0.001; **Figure 3A**). Linear regression showed a lack of proportional bias (β=- 0.026, SE=0.032, p=0.416) and Bland-Altman analysis resulted in limits of agreement between - 8.4 and 3.4 kg, with a maximum allowed difference ranging from -34.0 to 34.0 kg (**Figure 3B**). FM% measured with SF-BIA correlated strongly with DXA (r=0.867, p<0.001), with a lack of proportional bias (β=-0.003, SE=0.066, p=0.967) (**Figure 3C**), limits of agreement between -8.4 and 3.4%, and a maximum allowed difference from -16.7 to 16.7%) (**Figure 3D**).

**Figure 3.**
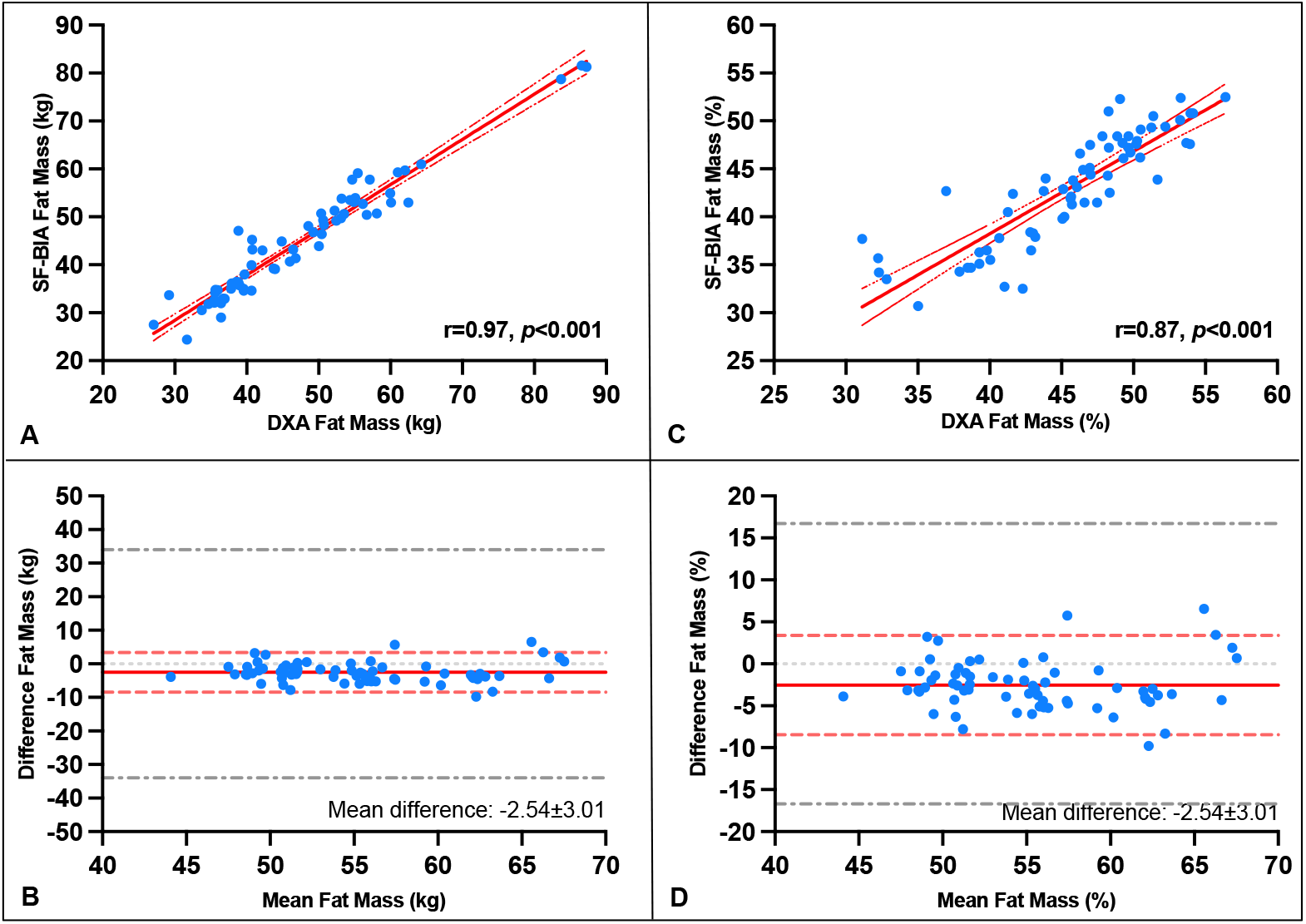
(A) Correlation of fat mass (FM) (kg) between DXA and SF-BIA measurement. (B) Bland-Altman plot with limits of agreement and maximum allowed difference of FM (kg) between SF-BIA and DXA measurement. (C) Correlation of fat mass (FM)% between DXA and SF-BIA measurement. (D) Bland-Altman plot with limits of agreement and maximum allowed difference of FM% between SF-BIA and DXA measurement. Dotted gray lines indicates maximum allowed difference; dotted red lines indicate limits of agreements; solid red line indicates mean difference between SF-BIA and DXA measurements.

Similarly, FFM (kg) measured by SF-BIA strongly correlated with that by DXA (r=0.954, p<0.001), and regression analysis showed no evidence of proportional bias (β=-0.052, SE=0.04, p=0.194) (**Figure 4A**), with limits of agreement between -3.5 and 10.2kg, and maximum allowed difference of (-30.2, 30.2 kg) (**Figure 4B**). For FFM%, the correlation was also strong (r=0.852, p<0.001), with no significant bias (β=-0.03, SE=0.071, p=0.669) (**Figure 4C**). Bland-Altman limits of agreement ranged between -2.5 and 9.3kg, and the maximum allowed difference from - 17.0 to 17.0 % (**Figure 4D**).

**Figure 4.**
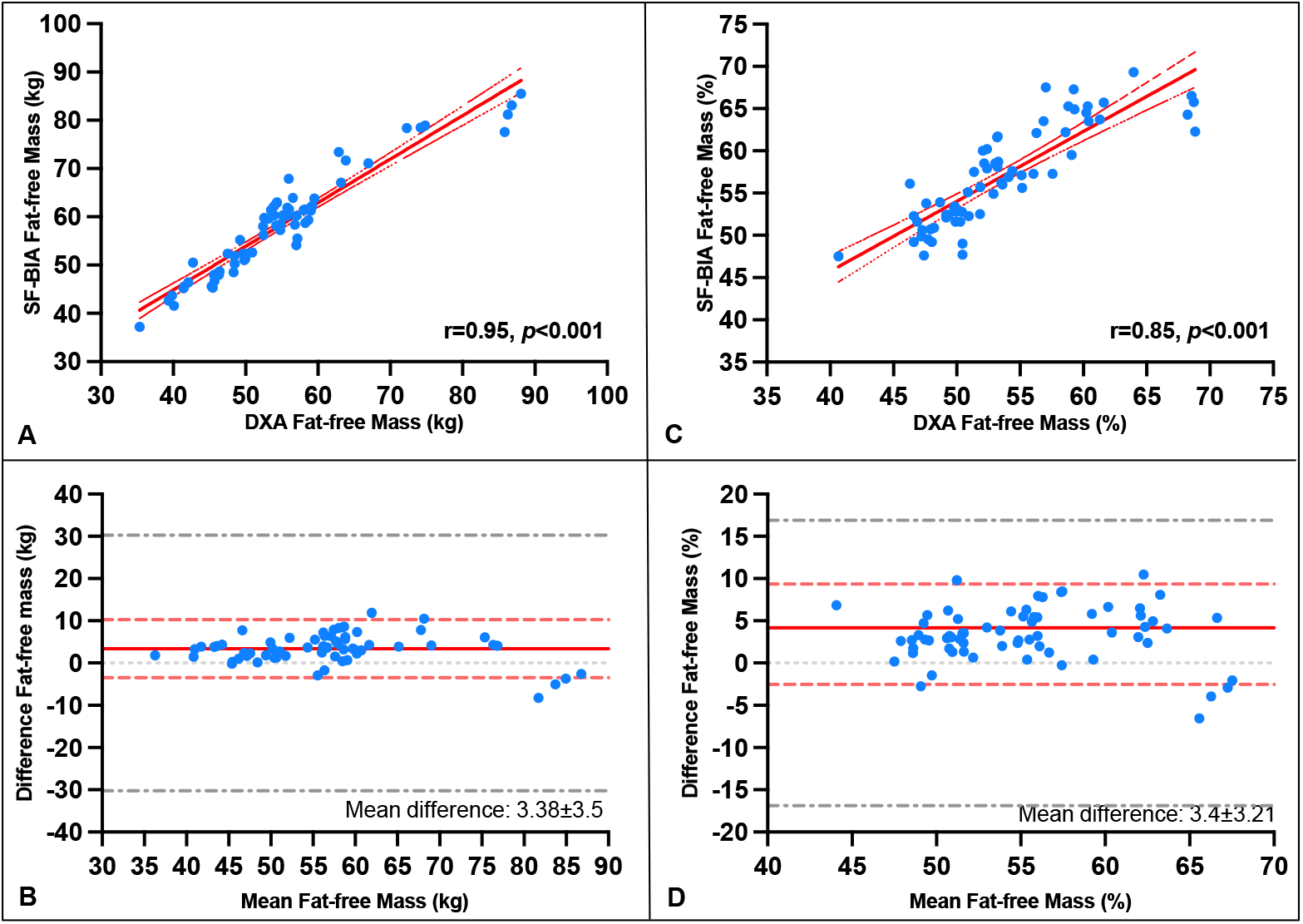
(A) Correlation of fat-free mass (FFM) (kg) between DXA and SF-BIA measurement. (B) Bland-Altman plot with limits of agreement and maximum allowed difference of FFM (kg) between SF-BIA and DXA measurement. (C) Correlation of fat-free mass (FFM) % between DXA and BIA measurement. (D) Bland-Altman plot with limits of agreement and maximum allowed difference of FFM % between SF-BIA and DXA measurement. Dotted gray lines indicates maximum allowed difference; dotted red lines indicate limits of agreements; solid red line indicates mean difference between SF-BIA and DXA measurements.

FFMI values measured by SF-BIA showed a strong, statistically significant correlation with DXA (r=0.88, p<0.001) (**Figure 5A**). Linear regression analysis indicated minimal bias (β=-0.013, SE=0.063, p=0.838), and Bland-Altman analysis displayed limits of agreement between -1.2 and 4.0 kg/m^2^, without any points outside the allowed range (-6.8, 6.8 kg/m^2^) (**Figure 5B**). FMI values also demonstrated a strong correlation (r=0.97, p<0.001) (**Figure 5C**). Regression analysis showed no evidence of proportional bias (β=-0.028, SE=0.078, p=0.721), and limits of agreement were calculated between -3.1 and 1.3 kg/m^2^, without outliers beyond the maximum allowed difference (-11.2, 11.2 kg/m^2^) (**Figure 5D**). **Table 2** summarizes the differences and related 95% confidence intervals between SF-BIA- and DXA-measured whole-body composition compartments.

**Figure 5.**
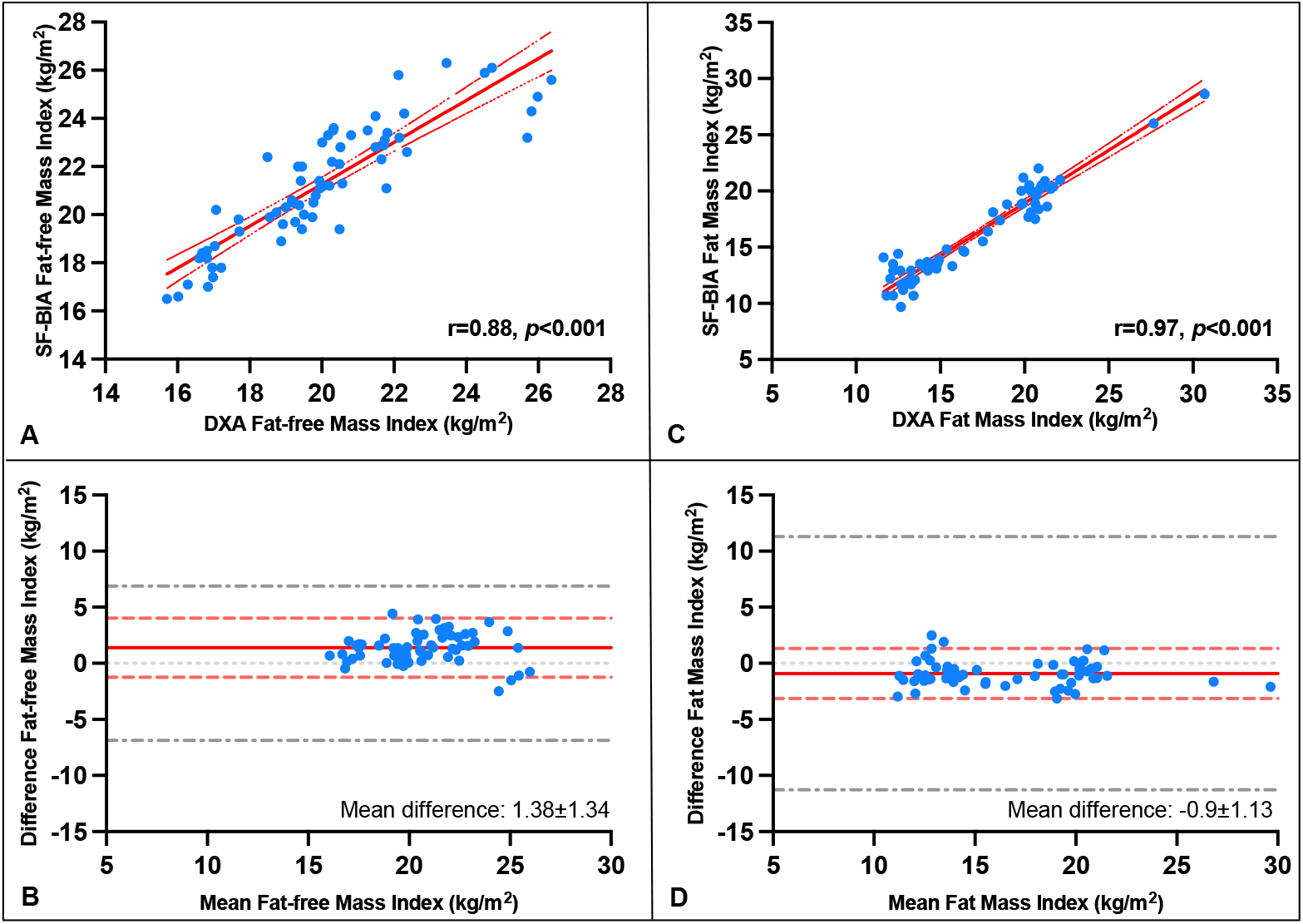
(A) Correlation of fat-free mass index (FFMI) (kg) between DXA and SF-BIA measurement. (B) Bland-Altman plot with limits of agreement and maximum allowed difference of FFMI (kg/m^2^) SF-BIA and DXA measurement. (C) Correlation of fat mass index (FMI) between DXA and SF-BIA measurement. (D) Bland-Altman plot with limits of agreement and maximum allowed difference of FMI between SF-BIA and DXA measurement. Dotted gray lines indicates maximum allowed difference; dotted red lines indicate limits of agreements; solid red line indicates mean difference between SF-BIA and DXA measurements.

## DISCUSSION

In this study, we found that segmental SF-BIA using the RJL Systems Quantum V device is a suitable alternative to DXA for measurement of whole- and segmental body composition in patients with obesity-related HFpEF. Except for AFMI, which is rarely utilized in research and clinical practice, all body composition compartments measured by the SF-BIA device presented a strong correlation without any evidence of proportional bias against DXA, a well-accepted reference standard tool to assess body composition. This is of particular clinical relevance because body composition plays a crucial role in determining CRF, quality of life, as well as survival in HF.^10, 21-26^ Moreover, up to 50% of patients with HF have a reduced ALMI, which portends to a worse prognosis.^26^ An accurate assessment of ALMI, a surrogate for skeletal muscle mass, also allows for the diagnosis of sarcopenia and sarcopenic obesity. Particularly, sarcopenic obesity has recently received intense scrutiny due to the pivotal role of weight loss in patients with obesity-related HFpEF. In fact, despite the short-term beneficial effects of weight loss in this population,^11, 27, 28^ FM loss is typically accompanied by a loss of skeletal muscle mass. Moreover, in patients undergoing energy restriction-induced weight loss, long-term weight regain in the form of FM is common, ultimately leading to a greater long-term risk for sarcopenic obesity.^21, 29^ In patients treated with medications for chronic weight management, weight regain is less common unless medication is interrupted; however, due to their powerful weight loss effects, whether the loss of skeletal muscle mass results in long-term detrimental effects is largely unknown.^30^ This is mainly due to the complexity of implementing available tools, such as DXA, resulting in the lack of body composition assessment in most pharmacologic trials in patients with obesity-related HFpEF.

In addition to providing an estimate of the quantity of body composition compartments, such as ALMI, this SF-BIA method also allows for the estimation of skeletal muscle quality in the form of phase angle, ultimately providing insights on both quantity and quality of body composition. Skeletal muscle quality is also a strong predictor for CRF assessed as VO_2 peak_ in patients with obesity-related HFpEF, however, its assessment remains challenging. We have previously reported that phase angle, purported to be a surrogate measure for cellular membrane integrity and body cell mass, is an independent predictor for VO_2 peak_ in patients with obesity-related HFpEF, where greater phase angle values were associated with more favorable CRF and exercise time.^31^

The results of this important study help identify alternative tools, filling an important and urgent gap in clinical and research practices. The RJL Systems Quantum V SF-BIA device offers a cost-effective and more accessible alternative to DXA for assessing body composition in patients with obesity-related HFpEF. Moreover, DXA uses the differential attenuation of two X-ray energies to perform multi-compartment assessments.^32^ The radiation dose from DXA is small (5-7μsv), but not insignificant, adding to the list of barriers to its broad implementation. Finally, the segmental body composition assessment with DXA usually requires a trained operator. In contrast to DXA, SF-BIA is radiation-free as it relies on differences of conductivity to assess body composition, passing a low voltage current between electrodes to measure body water, which is then used to estimate FFM, as well as FM.^19^ Moreover, SF-BIA requires only a few minutes of training since the segmental assessment is automatically estimated by the device and its related software. Also, SF-BIA can be safely utilized in individuals with implantable devices. The current findings support the use of the SF-BIA approach to measure body composition in HFpEF.

In conclusion, we propose that the segmental SF-BIA approach we evaluated is a practical, cost-effective, radiation-free alternative to DXA for accurately assessing whole-body, but most importantly segmental body composition in patients with persistent obesity-related HFpEF. As such, we advocate for its use in clinical trials, but also in clinical practice, to identify body composition phenotypes (i.e., sarcopenia, sarcopenic obesity) and to determine body composition compartments affected by a given intervention, whether it be lifestyle/behavioral or pharmacologic (i.e., medications for chronic weight management).

## Data Availability

Data are not being made available.

